## Supplementary material for "Suicide and self-harm in low- and middle- income countries during the COVID-19 pandemic: A systematic review"

**Summary of quality assessment criteria for reasonable quality studies**

| Study design | Assessment tool | Questions from the overall scale number | Criteria to be met |
|---|---|---|---|
| Cohort study | JBI | 1 | Two groups were similar and recruited from the same population |
| | | 2 | Exposures were measured similarly to assign exposure status |
| | | 6 | Were strategies to deal with confounding factors stated |
| | | 7 | Suicidal behaviour was measured in a valid and reliable way |
| | | 9 | Follow-up was complete or reasons for loss to follow up were described and explored |
| Before and After | NIH tool | 3 | All eligible participants that met the prespecified entry criteria were enrolled |
| | | 6 | Outcome measures were prespecified, clearly defined, valid, reliable, and assessed consistently across all study participants |
| | | 7 | The same approach/data source was used for the pre-covid measures of suicidal behaviour as those collected during the pandemic period |
| | | 8 | There were no differences in level of missing data pre and during the pandemic period |
| Time series | EPOC RoB Tool | 1 | Intervention independent of other changes |
| | | 2 | Intervention unlikely to affect data collection (low or unclear risk) |
| | | 5 | Missing outcome measures were unlikely to bias the results |
| Cross sectional | JBI | 1 | Criteria for inclusion was clearly defined |
| | | 2 | Study participants and setting were described in detail |
| | | 6 | Were strategies to deal with confounding factors stated |
| | | 7 | Suicidal behaviour was measured in a valid and reliable way |
| Case series | JBI | 1 | Criteria for inclusion was clearly defined |
| | | 3 | Valid methods were used for identification of the condition for all participants included in the case series |
| | | 4 | Consecutive cases were included |
| | | 6 | Clear reporting of demographics of the participants in the study |

JBI – Joanna Briggs Institute; NIH – National Institutes of Health; EPOC – (Cochrane) Effective Practice and Organisation of Care; RoB – risk of bias

**Number of cases of suicide and self-harm used to calculate rate ratios**

| Author (year) | Number of cases* by pandemic period | |
|---|---|---|
| | Pre- | During |
| Thongchuam (2021) | 1 | 7 |
| Fidanci (2021) | 187 | 31 |
| Eray (2021) | 23 | 11 |
| Acharya (2020) | 9 | 4 |
| Sengupta (2020) | 33 | 50 |
| Behara (2021) | 105 | 61 |
| Jhanwar (2020) | 3 | 11 |
| Shrestha (2020) | 38 | 55 |

* Cases were either suicide deaths or self-harm attempts
